# Geographical Disparities in HIV Prevalence among Men Who Have Sex with Men and People Who Inject Drugs in Nigeria

**DOI:** 10.1101/2020.01.09.20017103

**Authors:** Amobi Onovo, Abiye Kalaiwo, Moses Katbi, Otse Ogorry, Antoine Jaquet, Olivia Keiser

## Abstract

**Background:** Assessment of geographical heterogeneity of HIV among men who have sex with men (MSM) and people who inject drugs (PWID) can usefully inform targeted HIV prevention and care strategies. We aimed to measure HIV prevalence and identify hotspots of HIV infection among MSM and PWID in Nigeria.

**Methods:** We included all MSM and PWID accessing HIV testing services across seven prioritized states (Lagos, Nasarawa, Akwa Ibom, Cross Rivers, Rivers, Benue and the Federal Capital Territory) in three geographic regions (North Central, South South, South West) between Oct 1, 2016 and Sept 30, 2017. We extracted data from national testing registers, georeferenced all HIV test results aggregated at the level of Local Government Areas (LGAs), and calculated HIV prevalence. We calculated and compared HIV prevalence from our study to the integrated biological and behavioral surveillance survey (IBBSS) 2014 and used global spatial autocorrelation and hotspot analysis to highlight patterns of HIV infection, and to identify areas of significant clustering of HIV cases.

**Findings:** A positive HIV test was reported in 12.1% (95%CI 9.7-13.1) and 11.8% (95%CI 9.3-12.7) of the 26,423 MSMs and 9,474 PWIDs, respectively. Global spatial autocorrelation Moran’s I statistics revealed a clustered distribution of HIV infection among MSMs and PWIDs with a <5% and <1% likelihood that this clustered pattern could be due to chance respectively. Significant clusters of HIV infection (Getis-Ord-Gi* statistics) confined to the North Central, South-South regions were identified among MSM and PWID. Compared to the 2014 IBBSS our results suggest an increased HIV prevalence among PWID and a substantial decrease among MSM.

**Interpretation:** This study identified geographical areas to prioritize for control of HIV infection among MSM and PWID, thus demonstrating that geographical information system technology is a useful tool to inform public health planning for interventions for epidemic control of HIV infection.

**Funding:** Data used for this study was collected from Key Population program in Nigeria through PEPFAR/USAID. OK was funded by the Swiss National Science Foundation (grant no 163878).

## Introduction

Geographic variation in HIV prevalence and incidence has been demonstrated in many sub-Saharan African countries.^1-7^ According to the National HIV/AIDS and Reproductive Health Survey (NARHS, 2012), HIV prevalence in Nigeria among the general population was estimated at 3.4% in 2012 ranging from 0.6% in a South West location (Ekiti) to 15.2% in a South-South location (Rivers) (UNAIDS 2014). Much less is known about the spatial distribution of HIV infection in key populations (KP) such as men who have sex with men (MSM) and people who inject drugs (PWID), who have the highest risk of HIV.

Overall HIV prevalence in sub-Saharan Africa (SSA) is four times higher in MSM than in other men.^8^ The epidemic in west Africa is mainly heterosexually driven, but recent data suggest that sex between men may play a significant role in the spread of HIV. ^9,10^ HIV prevalence among MSM varies, with reported estimates over the past 3 years of 9.8% in Banjul, The Gambia ^11^, 18.0% in Abidjan, Côte d’Ivoire ^12^, 34.3% in Accra-Tema in Ghana ^13^, 34.9% in Abuja, Nigeria and up to 50.0% among MSM engaging in sex work in Abidjan, Côte d’Ivoire.^14^ These reports indicate that a substantial number of infections occur among MSM, many of whom also have sex with women.^15^ MSM across many countries in sub-Saharan Africa face stigma and discrimination.^16^ In Nigeria, the Same-Sex Marriage Prohibition law was passed by the Senate in 2011 and the law was implemented in January 2014.^17^ The new law criminalized same-sex practices, including prohibiting participation in organizations, service provision, or meetings that support gay people, and punishes attempts to enter civil unions or publicly show same sex relationships.^18^

About one third of the global HIV infections outside sub-Saharan Africa are related to PWID, and this accounts for a growing proportion of persons living with HIV. PWID have a 28 times higher risk of HIV compared with the general population.^19^ This risk arises particularly from sharing needles and injection equipment but is reinforced through criminalization, marginalization and poverty. Overall, it is estimated that 6.5% of people who inject drugs who live in central and West Africa have HIV.^20^ Prevalence ranges from 3.4% in Nigeria to 8.5% in Sierra Leone.^21^ However, Nigeria has the highest number of people who inject drugs in the region, estimated at 45,000 in 2017.^22, 23^ HIV prevalence among women who inject drugs is much higher than among men who inject drugs. For example, in Senegal HIV prevalence among women and men who inject drugs is 28% and 7%, respectively. ^24^

Previous studies, particularly in South Africa, Botswana and Uganda have focused on developing statistical models to predict the prevalence of HIV in KP. ^25-27^ These models included multiple risk factors (e.g., circumcision, distance to road and condom usage) and have shown that there can be considerable small-scale heterogeneity in HIV prevalence and risk behaviors. However, none of these models tested whether there is a spatial pattern of HIV infection among key populations and the size of the studies was limited. Currently, there is great interest amongst public health planners in whether this geographic variation of HIV could be used to increase the cost-effectiveness of interventions by focusing interventions towards areas of highest need. ^28,29,30^ We therefore aim to quantify the geographic variation of HIV prevalence among MSM and PWID by age and sex, by using georeferenced HIV Testing Services data aggregated at the level of Local Government Areas (LGAs). We aim to detect hotspots and compare HIV prevalence from our study to the integrated biological and behavioral surveillance survey (IBBSS) 2014. ^31^

## Methods

### Study Setting

Nigeria is organized into 36 federating states and the Federal Capital Territory (FCT) which hosts the national government. The states are further sub-divided into 774 Local Government Areas (LGAs). The study was conducted in 93 LGAs spread across seven prioritized states (Akwa Ibom, Rivers, Cross Rivers, Benue, Nasarawa, Lagos and the FCT); in three geographic regions North Central (NC), South South (SS) and the South West (SW). Under the national KP program, these states were prioritized for prevention and comprehensive HIV/AIDS treatment interventions due to the high population density and the high number of People Living with HIV (PLHIV).

### HIV Testing Modalities for KP

The national KP program consists of an integrated HIV prevention and treatment program that identifies HIV-positive KP in the community and links them to care and treatment at the LGA level. The program provides targeted HIV testing services to KP and their high-risk contacts. The following services are provided: (1) Index partner and/or index testing (also referred to as partner testing/partner notification) is an approach where exposed contacts of an HIV-positive index case are offered HIV testing. (2) KP are tested in mobile or temporary testing locations, such as community centers, schools, workplaces, hotels, clubs, tents and vans. (3) Voluntary Counselling and Testing (VCT) includes testing in VCT centers outside of a health facility (e.g. One-Stop-Shops (OSS). The OSS model for KP established safe spaces in the communities for HIV prevention and treatment interventions. It integrates differentiated strategies that optimize efficiency along the 90-90-90 cascade.^32^ HIV test results are aggregated by KP group and LGA where the test was done. In order to reduce double-counting of individuals and account for re-testers in a reporting period, tracking system such as “unique identifiers” are established and used to monitor the frequency of contact/outreach of KP over time. A unique ID is generated and assigned to a KP group before HIV testing commences, and results collected are entered into an electronic database. Data validation is conducted every quarter by running a query of all individual level KP data on HIV testing using the IDs to determine first time testers and repeat testers. All duplicated KP data within the reporting period are de-duplicated before data transmission.

### Study Population and statistical analyses

We analyzed aggregate-level data from index testing, mobile and VCT testing modalities by counting the number of KP who received HIV testing services via the PEPFAR/Nigeria KP program between October 1, 2016 and September 30, 2017 at the LGA level. All MSM and PWID aged ≥15 years with a documented HIV test result were included. MSM was defined as (1) self-identification as male and (2) report of oral or anal sex with a man in the prior 12 months. PWID was defined as a person who self-reported drug injection during the past 2 years. All KP groups were screened prior to enrollment by known MSM or PWID recruited as peer navigators on the national KP program to ensure clients were really members of the target population.

We georeferenced all MSM and PWID who assessed HIV testing services at the level of LGA and performed four analyses. First, we constructed maps of HIV prevalence among MSM and PWID. Second, we used Spatial Autocorrelation (Global Moran’s I) statistics to measure the degree, to which HIV prevalence is clustered, dispersed or randomly distributed. The expected value under the null hypothesis is that *“there is no pattern of HIV infection in selected LGAs”*. Moran’s I values range from -1 indicating perfect dispersion to +1 indicating perfect spatial clustering. Third, we used hotspot analysis in the ArcGIS software to calculate the Getis-Ord Gi* statistic for each LGA. The resultant z-score identified where LGAs with either high or low HIV prevalence cluster spatially. Each LGA is analyzed within the context of neighboring LGAs. The larger the z-score, the more intense the clustering of high prevalences, or hot spots. Similarly, smaller z-scores indicate clustering of low prevalences. To be a statistically significant hot spot, an LGA will have a high HIV prevalence and be surrounded by other LGAs with high HIV prevalence. Fourth, the calculated HIV prevalence from our study was compared to the HIV prevalence among MSM and PWID from IBBSS 2014.

The goal of the IBBSS is to obtain serological, behavioral and HIV service coverage data on key and vulnerable populations. The 2014 IBBSS was conducted by the National Agency for the Control of AIDS (NACA) and other stakeholders and used Respondent-driven sampling (RDS) for the selection of MSM and PWID, a modified form of snowball sampling to identify hard to reach populations. Members of the communities, NGOs working with the target populations, and key informants for each target group (recruited as seeds) assisted in the identification of various locations where the target groups could be found. Seeds were identified through the NGO networks that historically provide support and services for MSM and PWID. A list of sites where the population groups were located, how and when they can be reached for information and services and the essential distinguishing characteristics of these sites was prepared and used for the 2014 IBBSS. All eligible key and vulnerable populations were tested for HIV, and results were documented in survey forms.

### Ethical review

This analysis was conducted with routine data gathered through the national KP program. Informed consent was obtained for all clients who were tested for HIV in line with the Nigeria HTS policy. Ethical approval was obtained from the Federal Capital Territory, Health Research Ethics Committee, Nigeria (Approval Number: FHREC/2019/01/122/23-12-19). This study only analyzed anonymized and de-identified data.

## Results

### Overall HIV prevalence by age and sex

Of the 26,423 MSMs and 9,474 PWID who received HIV testing between October 1, 2016 and September 30, 2017, 3,209 MSM and 1,126 PWID tested HIV positive for an overall prevalence of 12.1% (95%CI 9.7-13.1) and 11.8% (95%CI 9.3-12.7) (<u>Table 1</u>). MSM aged 50 years and older had a considerable higher HIV prevalence (34.1%) compared to those aged 15-19 years (7.2%). Middle-aged PWID (25-49 years old) had the highest HIV prevalence (14.6%, n=6,203) compared to other PWID age groups. HIV prevalence among female PWID was twice higher than in male PWID (18.8% vs. 9.4%). Female PWID aged 25-49 years had the highest HIV prevalence (24.7%), while the prevalence among male PWID in the same age group was 11.7% (<u>Table 1</u>).

**Table 1:** HIV Prevalence among men having sex with men (MSM) and people who inject drugs (PWID) by age, sex and study locations

| State | Age | Number of individuals who received HIV Testing Services and received their test result |  |  | Number of individuals who tested HIV positive |  |  | HIV Prevalence among MSM | HIV Prevalence among Male PWID (%) | HIV Prevalence among Female PWID (%) |
| --- | --- | --- | --- | --- | --- | --- | --- | --- | --- | --- |
|  |  | MSM | PWID |  | MSM | PWID |  | (%) | (%) | (%) |
|  |  |  | Male | Female |  | Male | Female |  |  |  |
| Akwa Ibom State | 15-19 | 227 | 75 | 29 | 25 | 2 | 3 | 11.0% | 2.7% | 10.3% |
|  | 20-24 | 1134 | 281 | 82 | 53 | 13 | 20 | 4.7% | 4.6% | 24.4% |
|  | 25-49 | 2242 | 1033 | 188 | 490 | 159 | 66 | 21.9% | 15.4% | 35.1% |
|  | 50+ | 32 | 13 | 3 | 13 | 4 | 2 | 40.6% | 30.8% | 66.7% |
| — |  |  |  |  |  |  |  |  |  |  |
| Benue State | 15-19 | 264 | 42 | 14 | 16 | 6 | 3 | 6.1% | 14.3% | 21.4% |
|  | 20-24 | 1093 | 251 | 66 | 70 | 10 | 10 | 6.4% | 4.0% | 15.2% |
|  | 25-49 | 1117 | 875 | 117 | 282 | 85 | 40 | 25.2% | 9.7% | 34.2% |
|  | 50+ | 4 | 31 | 5 | 4 | 6 | 1 | 100.0% | 19.4% | 20.0% |
| — |  |  |  |  |  |  |  |  |  |  |
| Cross River State | 15-19 | 327 | 100 | 64 | 30 | 3 | 11 | 9.2% | 3.0% | 17.2% |
|  | 20-24 | 1495 | 358 | 176 | 145 | 11 | 27 | 9.7% | 3.1% | 15.3% |
|  | 25-49 | 2193 | 946 | 344 | 349 | 149 | 122 | 15.9% | 15.8% | 35.5% |
|  | 50+ | 31 | 9 | 1 | 11 | 4 | 0 | 35.5% | 44.4% | 0.0% |
| — |  |  |  |  |  |  |  |  |  |  |
| Federal Capital Territory | 15-19 | 753 | 28 | 41 | 21 | 0 | 0 | 2.8% | 0.0% | 0.0% |
|  | 20-24 | 2401 | 318 | 280 | 154 | 6 | 19 | 6.4% | 1.9% | 6.8% |
|  | 25-49 | 2332 | 677 | 293 | 243 | 24 | 26 | 10.4% | 3.5% | 8.9% |
|  | 50+ | 10 | 2 | 1 | 5 | 1 | 1 | 50.0% | 50.0% | 100.0% |
| — |  |  |  |  |  |  |  |  |  | — |
| Lagos State | 15-19 | 496 | 6 | 23 | 34 | 0 | 5 | 6.9% | 0.0% | 21.7% |
|  | 20-24 | 1375 | 44 | 76 | 167 | 2 | 5 | 12.1% | 4.5% | 6.6% |
|  | 25-49 | 731 | 278 | 219 | 99 | 4 | 26 | 13.5% | 1.4% | 11.9% |
|  | 50+ | 10 | 96 | 23 | 2 | 1 | 1 | 20.0% | 1.0% | 4.3% |
| — |  |  |  |  |  |  |  |  |  |  |
| Nasarawa State | 15-19 | 227 | 51 | 21 | 7 | 1 | 1 | 3.1% | 2.0% | 4.8% |
|  | 20-24 | 1294 | 369 | 181 | 97 | 8 | 13 | 7.5% | 2.2% | 7.2% |
|  | 25-49 | 1193 | 668 | 238 | 153 | 68 | 65 | 12.8% | 10.2% | 27.3% |
|  | 50+ | 6 | 3 | 0 | 2 | 1 | 0 | 33.3% | 33.3% | — |
| — |  |  |  |  |  |  |  |  |  |  |
| Rivers State | 15-19 | 354 | 7 | 0 | 57 | 2 | 0 | 16.1% | 28.6% | — |
|  | 20-24 | 1929 | 88 | 0 | 268 | 15 | 0 | 13.9% | 17.0% | — |
|  | 25-49 | 3120 | 327 | 0 | 406 | 71 | 0 | 13.0% | 21.7% | – |
|  | 50+ | 33 | 13 | 0 | 6 | 3 | 0 | 18.2% | 23.1% | – |
| Overall | 15-19 | 2,648 | 309 | 192 | 190 | 14 | 23 | 7.2% | 4.5% | 12.0% |
|  | 20-24 | 10,721 | 1,709 | 861 | 954 | 65 | 94 | 8.9% | 3.8% | 10.9% |
|  | 25-49 | 12,928 | 4,804 | 1,399 | 2,022 | 560 | 345 | 15.6% | 11.7% | 24.7% |
|  | 50+ | 126 | 167 | 33 | 43 | 20 | 5 | 34.1% | 12.0% | 15.2% |
| Total |  | 26,423 | 6,989 | 2,485 | 3,209 | 659 | 467 | 12.1% | 9.4% | 18.8% |

### HIV prevalence by state: comparison to previous surveys

<u>Figure 1a</u> shows median HIV prevalence for the different states. Among MSM in all states, the median HIV prevalence in the FCT was the highest (20.0%, 95%CI 3.4-25.9), followed by Lagos (13.5%, 95%CI 10.6-18.2) and Akwa Ibom state (12.0%, 95%CI 10.2-16.9). Median HIV prevalence was lowest in Nasarawa (2.0%, 95%CI 1.0-14.9). Similarly, among PWID, median HIV prevalence in the FCT was the highest 19.0% (95%CI 7.5-24.5), followed by Lagos (18.5%, 95%CI 9.5-19.3) and Nasarawa (13.0, 95%CI 4.1-18-8). Median HIV prevalence among PWID was the lowest in Benue (3.0%, 95%CI 0.6 – 12.7) (Figure 1a).

**Figure 1:**
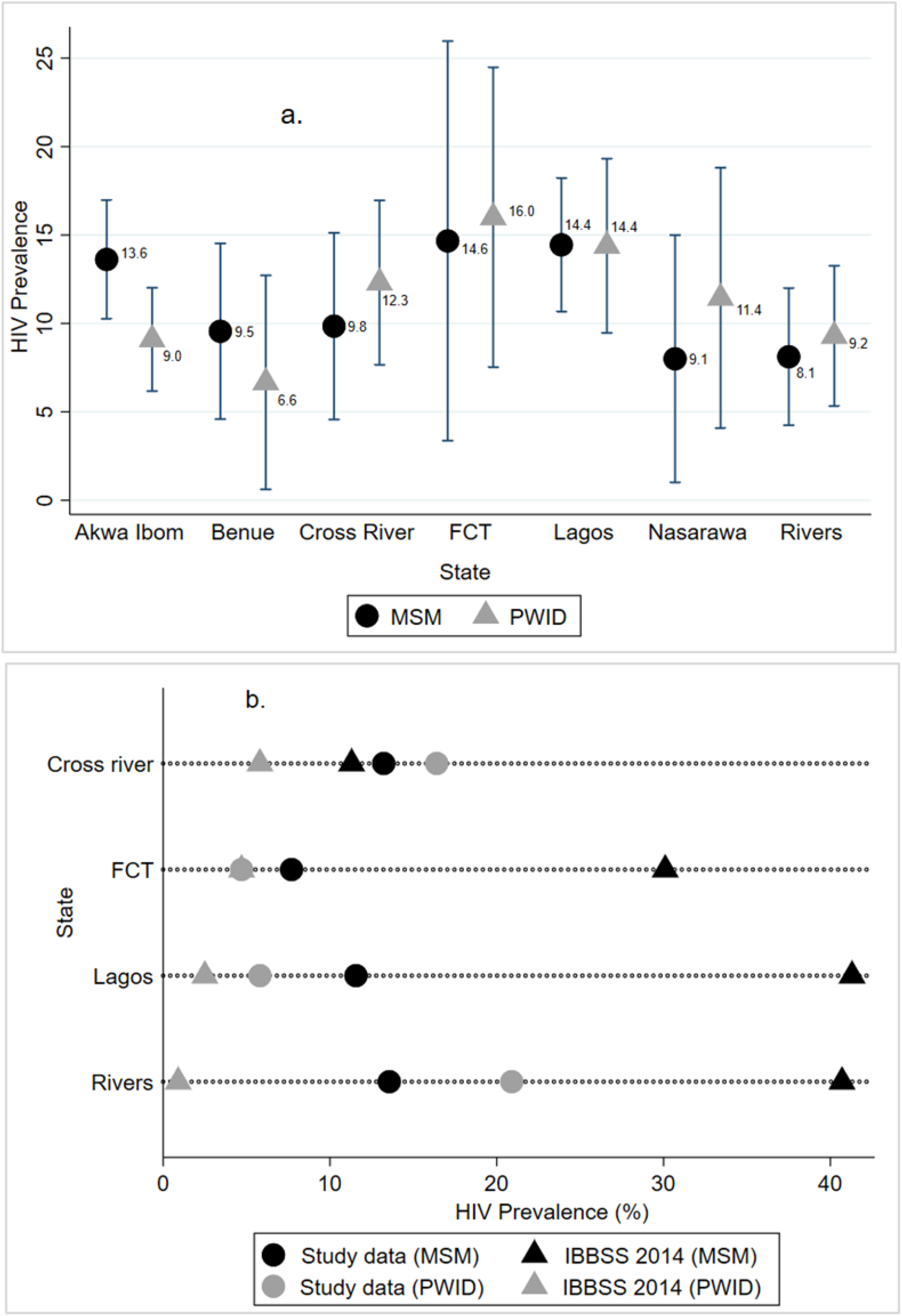
HIV prevalence among men who have sex with men (MSM) and people who inject drugs (PWID) by state. **a:** Mean prevalence with 95% confidence intervals. **b:** Comparison of HIV prevalence from the study to HIV prevalence in IBBS 2014

<u>Figure 1b</u> shows HIV prevalence for the different states in comparison to previous estimates from the 2014 IBBSS. Findings from the 2014 IBBSS have shown that HIV prevalence increased among MSM: from 13.5% in 2007, 17.2% in 2010 to 23.0% in 2014. While the prevalence among PWID decreased from 5.6% in 2007, 4.2% in 2010, to 3.4% in 2014. The IBBSS of 2014 was conducted in 14 states, and only 4 of the 7 states in our study (i.e. Lagos, Rivers, FCT and Cross River) had data on prevalence for MSM and PWID recorded in the IBBSS 2014 report. The graph shows clear differences in prevalence between IBBSS 2014 and our program data for MSM in Lagos (41.3% vs. 11.6%), Rivers (40.7% vs. 13.6%) and the FCT (30.1% vs. 7.7%). Prevalence from program data was higher than prevalence from IBBSS 2014 for MSM in Cross River state (13.2% vs. 11.3%) (<u>Figure 1b</u>). Prevalence among PWID was higher in our program data compared to IBBSS 2014 data in Lagos (5.8% vs. 2.5%), Rivers (20.9% vs. 0.9%) and Cross River (16.4% vs. 5.8%). Noteworthy to mention, HIV prevalence of the general population in the FCT (7.5%) based on NARHS 2012 was higher than the prevalence of PWID from program data (4.7%) and IBBSS 2014 (4.7%) in the same state (<u>Figure 1b</u>).

### Variability of HIV prevalence within different states

The interpolated HIV prevalence map reveals the geospatial distribution of HIV prevalence among MSM (<u>Figure 2a: top panel</u>) and PWID (<u>Figure 2b: bottom panel</u>) within different states. Prevalence among MSM was higher than in PWID. Large-scale spatial patterns are apparent, and they are more distinct for MSM than for PWID. Among MSM in Benue state, a high prevalence (>13%) was clear in Logo community, while in Gwer East and Vandeikya LGAs the prevalence was below 10% (Annex 2, <u>Figure 3a</u>). HIV prevalence among MSM in Rivers state was highest in Asari-toru, Emohua, Ahoada East, Ahoada West and Abua/Odual LGAs (<u>Figure 3b</u>). Similarly, HIV prevalence was relatively high in Rivers among PWID who live outside the mapped urban area settlements in Ahoada East, Ahoada West, Emohua, Abua/Odual and Bonny (<u>Figure 3c</u>). Notably, in Rivers state, high HIV prevalence among MSM was recorded in large urban area settlements in Asari-Toru LGA, and prevalence in Cross river among PWID was highest in urban area settlements in the northern region of Obubra LGA (<u>Figure 3d</u>).

**Figure 2:**
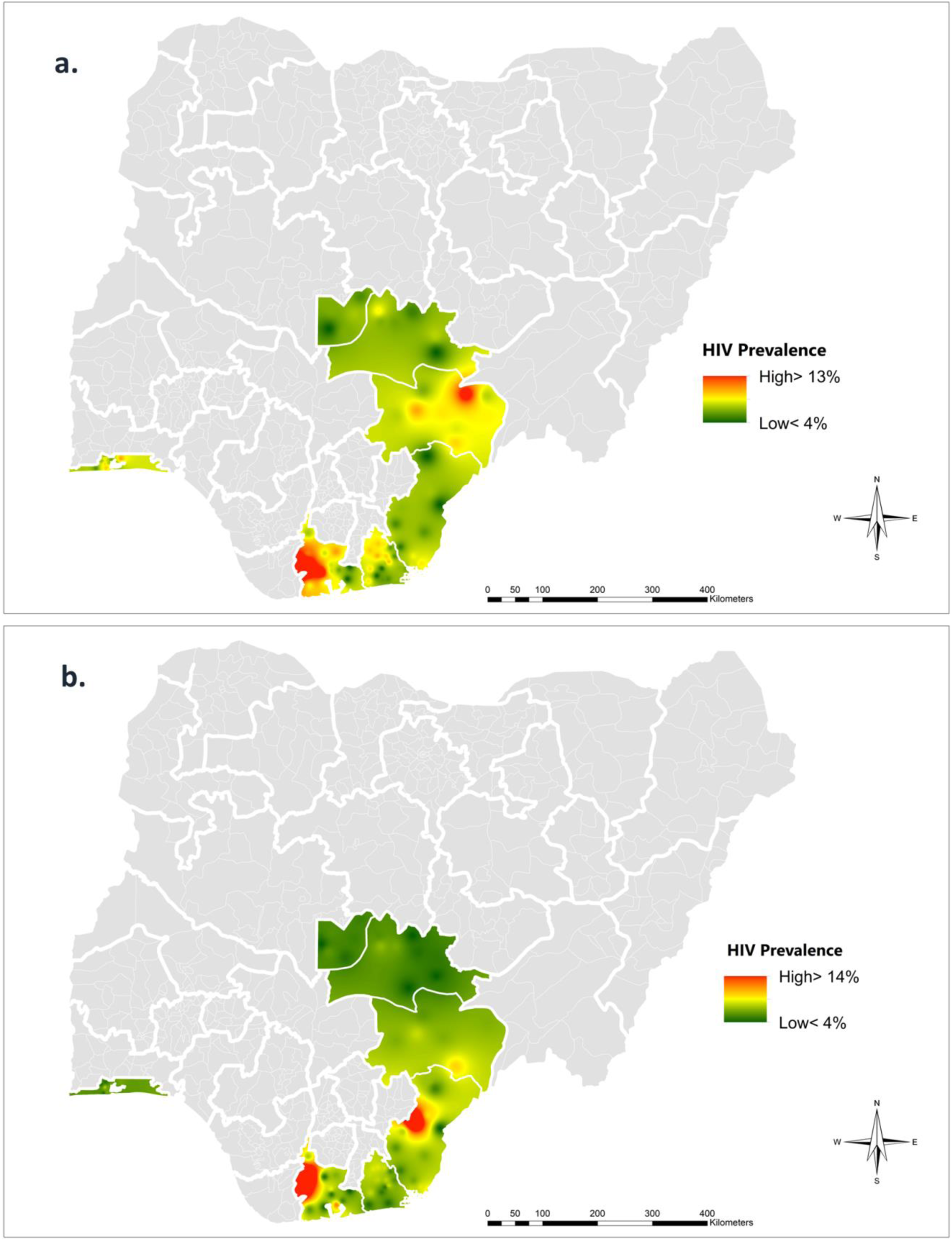
Map of Nigeria showing the interpolated areas with high and low HIV prevalence among men having sex with men (MSM) (a. top panel), and people who inject drugs (PWID) (b. bottom panel). Prevalence >13% are shown in red and prevalence <4% are shown in green.

**Figure 3:**
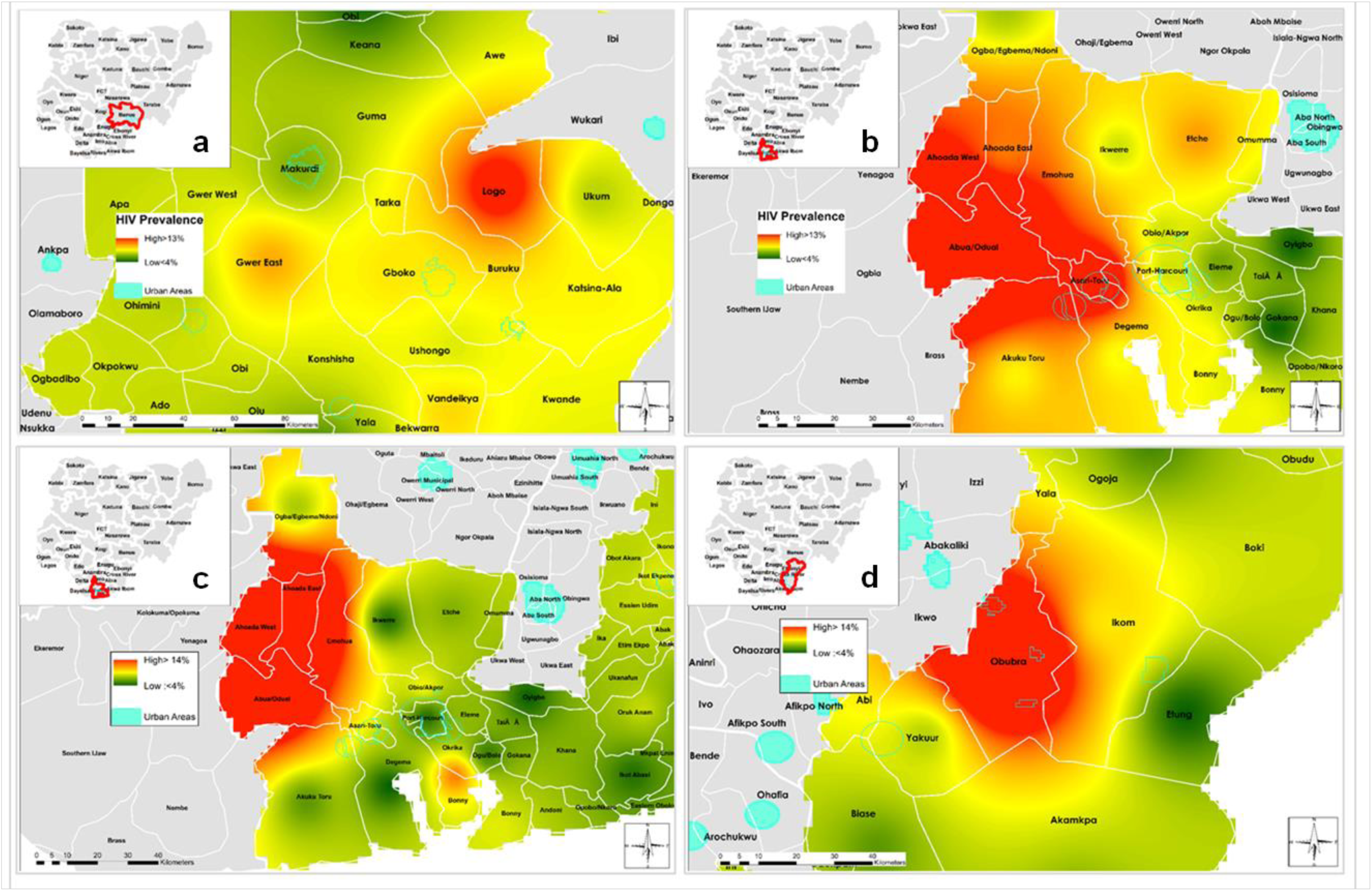
State level map showing urban locations and interpolated areas with high and low HIV prevalence by LGA among men having sex with men (MSM) (a. Benue and b. Rivers), and people who inject drugs (PWID) (c. Rivers and d. Cross River). Prevalence >13% are shown in red and prevalence <4% are shown in green.

### Detection of spatial patterns and hotspots of HIV prevalence

The global spatial autocorrelation Moran’s I statistics confirmed the clustered distribution of HIV prevalence among MSM and PWID. We found more significant clustering among PWID (z score=4.03, p<0.001) compared to MSM (z-score=2.29, p< 0.021). Getis-Ord-Gi* statistics indicated significant clusters of HIV infection among MSMs and PWIDs (<u>Table 2</u>) that were confined to LGAs in the NC (AMAC, Bwari, Karu, Keffi and Gwer East) and SS (Calabar Municipal, Calabar South, Degema, Odukpani and Akpabuyo) regions of the country.

**Table 2:** Hot spot analysis (Getis-Ord-Gi*) of HIV infection distribution among men having sex with men (MSM) (a. left) and people who inject drugs (PWID) (b. right) Only LGAs with a p value < 0.05 are shown

| A. |  |  |  |  | B. |  |  |  |  |
| --- | --- | --- | --- | --- | --- | --- | --- | --- | --- |
| LGA | MSM (#<br>HIV<br>Positive) | GiZScore | GiPvalue | GiBIN | LGA | PWID (#<br>HIV<br>Positive) | GiZScore | GiPvalue | GiBIN |
| Calabar<br>Municipal | 234 | 1.977624 | 0.047971 | 2 | Gwer East | 71 | 1.897651 | 0.057742 | 1 |
| AMAC | 430 | 3.282569 | 0.001029 | 3 | Calabar<br>South | 97 | 4.007088 | 0.000061 | 3 |
| Bwari | 69 | 4.205538 | 0.000026 | 3 | Calabar<br>Municipal | 157 | 4.78932 | 0.000002 | 3 |
| Karu | 155 | 3.559387 | 0.000372 | 3 | Obubra | 3 | 2.511684 | 0.012016 | 2 |
| Degema | 58 | 2.157567 | 0.030962 | 2 | Keffi | 104 | 2.046563 | 0.040701 | 2 |
| Port-Harcourt | 204 | 2.645243 | 0.008163 | 3 | Bakassi | 100 | 2.92129 | 0.003486 | 3 |
|  |  |  |  |  | Akpabuyo | 94 | 4.990284 | 0.000001 | 3 |

## Discussion

Our study shows that there is substantial geographic variation in the HIV prevalence of both MSM and PWID throughout the study areas. The interpolated maps in our analysis identified significant clustered patterns of the spatial distribution of HIV prevalence in Nigeria that would have been missed using macro-level national data. Overall, HIV prevalence among MSM ranged from 9.6% to 13.1% and among PWID from 9.3% to 12.7%. Prevalence was disproportionately distributed across the states. The median prevalence of 20.0% (95% CI: 1.0 – 24.0) and 19.0% (95% CI: 2.2– 23.0) for MSM and PWID in the FCT, and 18.5% (95%CI: 3.0 – 23.0) for PWID in Lagos shows a concentrated epidemic among these key populations in Nigeria. Surveillance data have shown that women carry the highest burden of HIV on the continent, with national-level statistics reporting that women have a higher HIV prevalence and incidence than men. ^33,34^ These reports are substantiated by findings from our study, which showed that HIV prevalence among female PWID was twice higher than in male PWID (18.8% vs. 9.4%).

Among MSM, the substantial decrease in HIV prevalence as observed in our study data compared to IBBSS 2014 could be attributed to prolonged exposure and effectiveness of HIV interventions in the study states over a 4-year period. The minimum comprehensive package of HIV interventions implemented in these states and targeted at the general population and key population groups included community and facility testing for HIV, linkage of HIV positive individuals to antiretroviral treatment and monitoring of HIV treatment retention to improve viral suppression. These finding from our study, are substantiated by reports from the Nigeria AIDS Indicator and Impact Survey (NAIIS) 2018 which showed a national HIV prevalence in Nigeria of 1.4% among adults aged 15–49 years compared to previous national prevalence estimates of 3.4% from the 2012 National HIV & AIDS and Reproductive Health Survey (NARHS Plus). NAIIS also showed that Nigeria has shown steady progress on increasing access to treatment for people living with HIV, with the adoption of a test and treat policy in 2016. Among PWID, our results indicated a higher prevalence compared to the 2014 IBBSS. The disproportionate high prevalence among PWID in our study compared to 2014 IBBSS could be linked to the HIV infection distribution pattern among PWID that suggests overlapping risk groups with multiple transmission routes. For example, some PWID are sex workers, or buy or trade drugs for sex, or are MSM, and may gain HIV through sexual and injecting routes.^34^

Recent studies outside sub-Saharan Africa has established high prevalence of HIV among overlapping high-risk populations such as people who inject drugs, men who have sex with men, and female sex workers, where there are multiple transmission routes. Epidemiological studies across 2003–2015 in Tijuana reported a pooled HIV prevalence of 3.4% among male PWID, but 8% among male PWID who have sex with men, and 5% among male PWID who are clients of FSW.^35^ Similarly, pooled studies from 2003–2015 reported 5% HIV prevalence among FSW, rising to 6.7% among female PWID, and 7.3% among FSW who inject drugs.^35^ In Pakistan, HIV is concentrated largely among PWID, with an estimated prevalence of 11.0% in 2004, increasing steadily to 27.2% by 2011.^36^ Systematic reviews and data synthesis from the middle east and north Africa found evidence of HIV prevalence among PWID overall in the range of 10%-15%, and highest HIV prevalence among PWID globally (87.1% in Tripoli, Libya).^37^ The relatively high prevalence of sharing needles/syringes (18%-28% in the last injection), the low levels of condom use (20%-54% ever condom use), the high levels of having sex with sex workers and of men having sex with men (15%-30% and 2%-10% in the last year, respectively), and of selling sex (5%-29% in the last year), indicate a high injecting and sexual risk environment.^37^ The 2018 National Survey on Drug Use and Health in Nigeria reported that People who inject drugs constitute a sizeable proportion of high risk drug users in Nigeria. 1 in 5 high risk drug users is injecting drugs. The most common drugs injected in the past year were pharmaceutical opioids, followed by cocaine and heroin. Women who injected drugs were more likely than men to engage in high-risk sexual behaviors further compounding their risk for acquiring HIV among other infections.^38^ These results further collaborate with findings from our study that showed high prevalence of HIV among female PWID than male PWID.

A key limitation of our study was that criminalization through the Same Sex Marriage Prohibition Act, and discrimination against KPs has driven several KPs underground, limiting the overall representation of the study. Consequently, continued discrimination and criminalization of KPs impede access to health services and willingness to participate in surveys. In 2014, the Nigerian government increased the punishment for homosexuality to 14 years in jail. Anyone who assists homosexual couples may face up to 10 years in prison.^39^ Mass arrests of suspected gay men in Nigeria have followed, for example in July 2017 the police arrested 40 men at a private house party.^40^ Criminalizing laws such as these have pushed MSMs into hiding, making them more vulnerable to HIV.^41^ Program data used for this analysis was collected at aggregate level, and did not include socio-economic and/or clinical characteristics of the study population that might have provided further insights on the variation of HIV seropositivity.

The large sample of KP data collected and used in our study compared to other previous studies can be attributed to the peer outreach model. In this model, KPs are trained as peer-outreach workers to increase demand for tailored HIV services, improve the quality of behavior change communication and increase access to HIV testing services, the starting point of the key population cascade, via social networks. This approach yielded representative data about the target KP groups at the LGA level and offers a unique opportunity to estimate national and subnational key population size through scaling up the use of the peer outreach method for data collection in HIV programming. The large sample of MSM and PWID data as reflected in our study is an indication that KP can be effectively mobilized for HIV testing and treatment despite legal, policy and social barriers.

## Conclusions

In conclusion, we have provided, to our knowledge, the largest study on HIV prevalence in KP in SSA and we showed that there is substantial clustering and subnational variation in HIV prevalence among MSM and PWID. Our results suggest that HIV prevalence among middle-aged adult female PWID is disproportionately greater than among male PWID. PWID could therefore transmit their HIV infection to their sexual contacts and injecting partners and this in turn may lead to the spread to the general population, particularly in the context of transactional sex. Interventions that significantly focus on women and their sexual and injecting partners and/or clients are of importance to address these high HIV acquisition and transmission risks among KP, in particular PWID. The substantial decrease in HIV prevalence among MSM as observed in our study data compared to IBBSS 2014 suggests improvements in the national HIV prevention and treatment programmes in the past four-years. Understanding heterogeneity in mixing patterns among KPs in concentrated HIV epidemics may help design more effective interventions. We recommend the use of routine HTS program data for implementation of HIV infection surveillance among KP. They should serve as an essential input for statistical models that estimate the national and subnational burden and incidence of HIV using estimation and projections tools.

## Data Availability

Data used for this study was collected from Key Population program in Nigeria through PEPFAR/USAID. This study only analyzed anonymized and de-identified data. All the data included for analysis in this study is available.

## Contributors

AAO conceived of the study including design and method. He was the principal in data management, did the data analysis and wrote the manuscript draft. OK performed critical reviews of the first and final version of the manuscript and provided input into the analysis and manuscript. AK, MK, UR, and OO provided reviews and feedback at the abstract phase. AJ reviewed and provided feedback on the final version of the manuscript. All authors approved the final version of the manuscript.

## Declaration of interests

In this study, we report no financial or non-financial competing interests.

## Acknowledgments and funding

Data used for this study was collected from Key Population program in Nigeria through PEPFAR/USAID. OK was supported by a professorship grant from the Swiss National Science Foundation (grant no 163878). We thank staff of Heartland Alliance Nigeria for their contribution on data collection and data transmission from program implementation states. We thank Ben Spycher for comments on an earlier version of this manuscript.

**Research in Context**

**Evidence before this study**

Nigeria’s HIV situation is a complex, mixed epidemic, driven by diverse factors depicting substantial differences in HIV prevalence across different regions and sub-populations. Currently, there is great interest among public health planners in whether the geographic disparity in HIV prevalence could be used to increase the cost-effectiveness of HIV/AIDS interventions by focusing interventions towards areas of highest need. The present study focuses on MSM and PWID because of their heightened risk of HIV acquisition and transmission. MSM across many geographical areas in Nigeria face stigma and discrimination. About one-third of the global HIV infections outside sub-Saharan Africa are related to People Who Inject Drugs (PWID), and this accounts for a growing proportion of persons living with HIV. We searched Google Scholar and PubMed from inception until October 30, 2018, for publications in English using the terms “HIV”, “Prevalence”, “MSM”, “PWID” and “Geospatial”, or “Disparity” and “Nigeria”. Our search returned over 17 reviews from PubMed and 8 reviews from Google Scholar. About 15% of the reviews reported that KPs are a hidden population, and reliable and representative epidemiological data on their health are scarce. Our search also found a study from Uganda, focusing on developing complex statistical models to predict the prevalence of HIV among KP. This model included multiple risk factors (e.g., circumcision and condom usage) and has shown that there can be considerable small-scale heterogeneity in HIV prevalence and risk behaviors.

**Added Value of this Study**

To the best of our knowledge, this is the first direct, ecological descriptive evidence of the spatial clustering of HIV prevalence among MSM and PWID in the national KP focus states in Nigeria. We found that HIV prevalence among middle-aged adult female PWID was twice higher than in male PWID. Similarities in the observed clustered distribution pattern of HIV infection among MSM and PWID suggest overlapping risk groups with multiple transmission routes. For example, according to recent publications, some PWID are sex workers, or buy or trade drugs for sex, or are MSM, and may gain HIV through sexual and injecting routes. We also highlight geostatistical methodologies that involved in the direct detection of spatial patterns and identification of hotspots.

**Implications of all the available evidence**

Underlying dynamics of the HIV infection distribution among MSM and PWID in our study show demographic and urban-centered characteristics may have contributed to the observed spatial clustering and disparities of the HIV infection. According to our study, HIV prevalence among men who have sex with men is decreasing compared to 2014 Biological and Behavioral Surveillance data that indicated otherwise. Our study suggests that inclusion of and significant focus on women and their clients are of importance to address these high HIV acquisition and transmission among KP, in particular PWID. Targeted, cost-effective programs that address not only behavioral but also biological and structural risk factors associated with HIV acquisition and transmission among key populations should be engaged to reduce the onward spread of HIV.

## Notes

### Competing Interest Statement

The authors have declared no competing interest.

### Funding Statement

No external funding was received. Authors were wholly responsible for the costs of the abstraction, communication, data collation, analysis and other associated costs.

